# Clinical and Economic Impact of Differential COVID-19 Vaccine Effectiveness in the United States

**DOI:** 10.1101/2022.03.31.22272957

**Authors:** Michael Maschio, Kelly Fust, Amy Lee, Nicolas Van de Velde, Philip O. Buck, Michele A. Kohli

## Abstract

**Background:** In the United States (US), three vaccines are currently available for primary vaccination and booster doses to prevent coronavirus disease 2019 (COVID-19), including the 2-dose messenger ribonucleic acid (mRNA) BNT162b2 (COMIRNATY®, Pfizer Inc) and mRNA-1273 (SPIKEVAX®, Moderna Inc) vaccines, which are preferred by the Centers for Disease Control and Prevention’s (CDC) Advisory Committee on Immunization Practice (ACIP), and the adenovirus vector Ad26.COV2.S (Johnson & Johnson) vaccine. A substantial body of evidence has now been published on the real-world effectiveness and waning of the primary series and booster doses against specific SARS-CoV2-variants. The study objective was to determine the clinical and economic impact of differences in effectiveness between mRNA-1273 and BNT162b2 booster vaccinations over one year (2022) in US adults ≥18 years.

**Methods:** A decision analytic model was used to compare three mRNA booster market share scenarios: (1) Current Scenario, where the booster mix observed in December 2021 continues throughout 2022; (2) mRNA-1273 Scenario, where the only booster administered in 2022 is mRNA-1273, and (3) BNT162b2 Scenario, where the only booster administered in 2022 is BNT162b2. Analyses were performed from the US healthcare system perspective. Sensitivity analyses were performed to explore the impact of COVID-19 incidence in the unvaccinated population and vaccine effectiveness (VE) on model results.

**Results:** In the Current Scenario, the model predicts 65.2 million outpatient visits, 3.4 million hospitalizations, and 636,100 deaths from COVID-19 in 2022. The mRNA-1273 Scenario reduced each of these outcomes compared to the Current Scenario. Specifically, 684,400 fewer outpatient visits, 48,700 fewer hospitalizations and 9,500 fewer deaths would be expected. Exclusive of vaccine costs, the mRNA-1273 Scenario is expected to decrease direct medical costs by $1.3 billion. Conversely, the BNT162b2 Scenario increased outcomes compared to the Current Scenario: specifically, 391,500 more outpatient visits, 34,500 more hospitalizations and 7,200 more deaths would be expected in 2022, costing an additional $946 million in direct medical costs. For both the mRNA-1273 and BNT162b2 booster scenarios, the percent change in direct treatment costs for COVID-19 is similar to the percent change in hospitalizations as the rate of hospitalizations is the driver of the overall costs.

Changing the number of projected COVID-19 cases in 2022 by varying the incidence rate has a direct effect on model outcomes. Higher incidence rates leads to higher outpatient visits, hospitalizations and deaths for all scenarios. Varying VE has an inverse effect on model outcomes. All outcomes increase when VE is lower for all vaccines and decrease when VE is higher. In all cases, additional use of mRNA-1273 leads to fewer infection outcomes while additional use of BNT126b2 results to higher infection outcomes.

**Conclusion:** As the real-world effectiveness evidence to date indicates that mRNA-1273 may be more effective at preventing COVID-19 infection and hospitalization over time than BNT-162b2, increasing the proportion of people receiving this as a booster are expected to reduce COVID-19-related outcomes and costs in 2022, regardless of COVID-19 incidence or variant.

## 1. INTRODUCTION

In the United States (US), there are currently three vaccines approved for active immunization to prevent coronavirus disease (COVID-19) and recommended for primary vaccination and booster doses by the Centers for Disease Control and Prevention’s (CDC) Advisory Committee on Immunization Practice (ACIP).^1^ The BNT162b2 (COVID-19 vaccine, COMIRNATY®; Pfizer Inc, New York, NY, USA; BioNTech Manufacturing GmbH, Mainz, Germany) and mRNA-1273 (COVID-19 Vaccine, SPIKEVAX®; Moderna Inc, Cambridge, MA, USA) vaccines are 2-dose messenger ribonucleic acid (mRNA) vaccines with full biologics license application (BLA) approval by the US Food and Drug Administration (FDA),^2,3^ and the Ad26.COV2.S (COVID-19 Vaccine; Johnson & Johnson, Beerse, Belgium) vaccine is a single-dose adenovirus vector-based vaccine authorized under emergency use authorization (EUA) by the FDA.^1,4^ Although all three vaccines are approved for use in the US, ACIP advises that the use of the mRNA vaccines are preferred due to lower effectiveness and risk of serious adverse events.^5,6^

ACIP defines a booster dose as a “dose of vaccine administered to enhance or restore protection by the primary vaccination, which might have waned over time” and recommends any of the three COVID-19 vaccines, regardless of type of vaccine received in the primary series, may be used for the purposes of a booster dose.^1^ ACIP recommends that all adults, ages 18 and older, receive a COVID-19 booster shot.^7^ Per ACIP, people who received either of the mRNA vaccines for their primary series should receive a booster at least 5 months after the primary series is complete, while those who received the Ad26.COV2.S primary vaccine should receive a booster dose at least 2 months following primary vaccination.^7,8^ ACIP also recommends that all people receive either one of the mRNA vaccines for the booster dose, regardless of primary vaccine type received, as these are preferred to the Ad26.COV2.S vaccine.^7^

Based on data provided by the CDC, as of the end of December 2021, approximately 68.7% of US adults ages 18-64 years and 87.5% of adults ≥65 years had received their primary series; of those receiving their primary series, 29.4% of those 18-64 years and 58.5% of those ≥65 years had also received a booster dose.^9^ The current scenario, as it pertains to boosting, includes a mixture of primary series and booster dose types. The majority of people who received either the BNT162b2 or mRNA-1273 primary series also received either the BNT162b2 or mRNA-1273 boosters (94.7% and 93.7%, respectively, of those receiving a booster), with the remainder receiving the Ad26.COV2.S booster.^10^ Of those who received the Ad26.COV2.S primary series, 27.2% also received the Ad26.COV2.S booster, 41.8% received the mRNA-1273 booster, and 31.1% received the BNT162b2 booster.^10^

There is a growing body of effectiveness data for these vaccines.^11-15^ While test-negative case controlled studies are the preferred method of estimating vaccine effectiveness (VE) using real world data,^16^ in the case of COVID-19, these data are often outdated by time of study completion as new variants emerge. Predictive models have also been developed to estimate VE over time by correlating levels of vaccine-induced antibodies against the variant of concern (neutralizing titers) to observed VE from clinical trials and real-world test-negative case-control study results.^17-19^ Thus, neutralizing titer levels may assist in the development of variant specific vaccines by allowing prediction of the level of VE over time with a moderate degree of certainty before real world data are available. Real world evidence and predictive modeling have shown that protection against symptomatic and severe infection vary by vaccine type, number of doses received, and timing since vaccination (waning). In addition, initial VE and rate of waning differs based on the circulating variants.^11,19-21^

While there is uncertainty as to how the COVID-19 pandemic will evolve, the current study uses an economic model to project various scenarios for 2022 with the aim of quantifying the potential impact of the differences in effectiveness between the two available mRNA booster vaccines. Three specific booster market share scenarios were examined, including the current scenario, which represents the mixture of booster doses across the three vaccine types as described above, and the hypothetical impact of changing the mix of boosters delivered to either 100% mRNA-1273 or 100% BNT162b2. The objective of this study was to estimate the clinical and economic impact of any potential differences in effectiveness between mRNA-1273 and BNT162b2 boosters in 2022 in adults aged 18 years and over in the US.

## 2. METHODS

### OVERVIEW

A decision analytic model was used to compare three mRNA booster market share scenarios: (1) Current Scenario, in which the mix of boosters administered by December 2021, which are presented in Table 1, remains constant throughout 2022; (2) mRNA-1273 Scenario, in which the only booster administered in 2022 is mRNA-1273 (i.e., mRNA-1273 booster has 100% market share starting in January), and (3) BNT162b2 Scenario, in which the only booster delivered in 2022 is BNT162b2 (i.e., BNT162b2 booster has 100% market share starting in January). The target population for the analysis consisted of adults aged 18 years and older in the US eligible for COVID-19 vaccination per ACIP recommendations.^1^ The time horizon of the analysis was 1 year (2022). Economic analyses were performed from the perspective of the US healthcare system and included direct healthcare costs only. Additional sensitivity analyses were performed to explore the impact of COVID-19 incidence in the unvaccinated population and VE on model results. In the model, the size of the US adult population and the number of patients receiving the primary series only and primary series with a booster dose were estimated, the number of infections in each group were calculated and a decision tree was used to estimate associated infection consequences and costs (Figure 1). The model was developed in Microsoft Excel using Visual Basic for Applications (Microsoft Corp; Redmond, Washington).

**Table 1.** Market share: vaccines received amongst those whose received the primary series and booster, by scenario^9,10^.

|  | Current Scenario | mRNA-1273 Scenario | BNT162b2 Scenario |
| --- | --- | --- | --- |
| **Primary Series (Ages 18-64 years)** |  |  |  |
| mRNA-1273 | 36.0% | 36.0% | 36.0% |
| BNT162b2 | 54.0% | 54.0% | 54.0% |
| Ad26.COVS2.S | 10.0% | 10.0% | 10.0% |
| **Primary Series (Ages ≥65 years)** |  |  |  |
| mRNA-1273 | 47.7% | 47.7% | 47.7% |
| BNT162b2 | 47.4% | 47.4% | 47.4% |
| Ad26.COVS2.S | 4.8% | 4.8% | 4.8% |
| **Booster Dose Received, Among Primary mRNA-1273 Recipients (Ages ≥18 years)** |  |  |  |
| mRNA-1273 | 93.7% | 100% | 0% |
| BNT162b2 | 6.2% | 0% | 100% |
| Ad26.COVS2.S | 0.1% | 0% | 0% |
| **Booster Dose Received, Among Primary BNT162b2 Recipients (Ages ≥18 years)** |  |  |  |
| mRNA-1273 | 5.3% | 100% | 0% |
| BNT162b2 | 94.7% | 0% | 100% |
| Ad26.COVS2.S | 0.0% | 0% | 0% |
| **Booster Dose Received, Among Primary Janssen/ Ad26.COVS2.S Recipients (Ages ≥18 years)** |  |  |  |
| mRNA-1273 | 41.8% | 100% | 0% |
| BNT162b2 | 31.1% | 0% | 100% |
| Ad26.COVS2.S | 27.1% | 0% | 0% |

**Figure 1.**
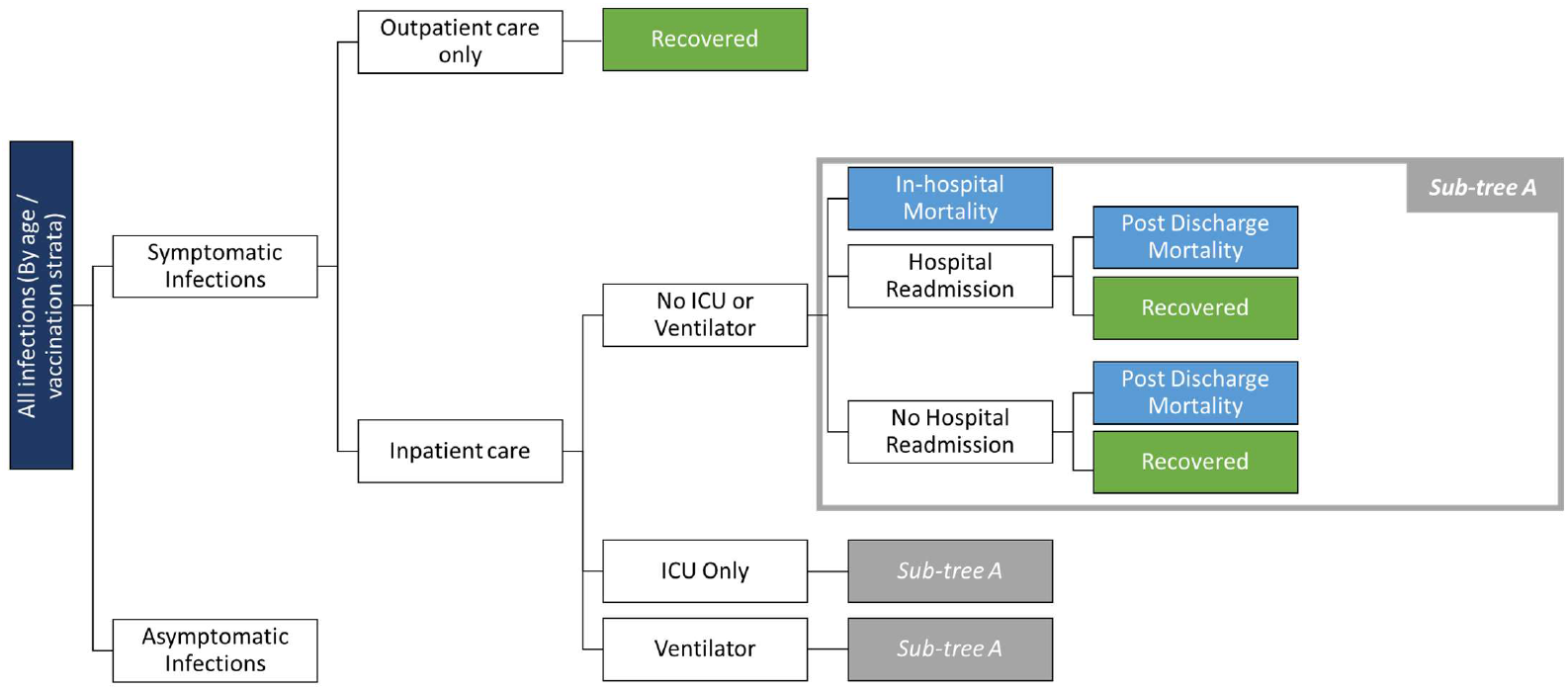
Infection Consequences Decision Tree.

### POPULATION, VACCINE COVERAGE, and MARKET SHARE

The size of the adult US population was estimated by age group using US Census Bureau data (See Appendix 2; Table 5).^22^ Primary series vaccine coverage was estimated using CDC data^9^ from March 2021 through December 2021 cumulatively at 68.7% for adults ages 18-64 and 87.5% ≥65 years. We made the simplifying assumption that no additional primary series were administered in 2022 as our analysis focuses on the impact of changes in booster delivery and primary series vaccinations have plateaued. December 2021 estimates of booster coverage among those who had previously received the primary series were also calculated from CDC data to be 29.4% in adults ages 18-64 and 58.5% in ≥65 years. Booster coverage was assumed to increase by 15% per month, until a maximum of 95% of those who had previously received the primary series was reached.

All market share estimates are displayed in Table 1. Values for the primary series reflect the distribution of the total number of patients receiving each vaccine as of the end of December 2021 for the 18-64 and ≥65 years age groups and was the same for all scenarios. The same data were also used to estimate the type of booster dose received. In the Current Scenario, primary series and booster market shares are assumed to be equivalent to the estimates at the model start for the duration of 2022. In the mRNA-1273 Scenario, 100% of booster doses are assumed to be mRNA-1273, and in the BNT162b2 Scenario, 100% of booster doses are assumed to be BNT162b2, regardless of primary series type received.

### DECISION TREE STRUCTURE

For each month of the analysis, the model groups people into unvaccinated, vaccinated with primary series only, and vaccinated with primary series plus booster using the coverage data described above. The risk of infection in the unvaccinated varies by month. Amongst those who received the primary series only or the primary series plus booster, the incidence of infection is reduced according to VE, and is recalculated each month to account for waning based on time since vaccination. For those who received the primary series in 2021, we made the simplifying assumption that their series was completed in May, as this marked the month at which 50% cumulative coverage in the population was achieved. May was therefore our anchor month to calculate time since primary series completion for the VE calculations for each month. The primary series VE was estimated as an average of the waning adjusted VE weighted by the proportion of the cohort receiving each primary series. For anyone who had received a booster in 2021, we made the simplifying assumption that the booster was received in November. As people continued to receive boosters in 2022, the booster VE was estimated as a weighted average of the waning adjusted vaccine effectiveness (time since receiving their booster) and the proportion of the cohort receiving each booster.

All patients who develop an infection are assumed to move into an infection consequences decision tree (Figure 1). Upon entry into the tree, infections are categorized as asymptomatic or symptomatic. Asymptomatic infections have no direct healthcare costs and are not modeled; accordingly, the model structure reflects symptomatic infections only. Patients with symptomatic infections may require either outpatient care only, or inpatient care. Those requiring outpatient care are subject to COVID-19 related healthcare costs and are assumed to survive the infection. As the vaccines have proven to be more effective at reducing severe disease than symptomatic disease alone, the model reduces the probability of hospitalization for patients who have received the primary series alone or the primary series plus booster using the VE data. Patients requiring inpatient care are distributed by location of care (no intensive care unit [ICU] or ventilator, ICU only, or ventilator). Hospitalized patients are assumed to incur inpatient costs and are at risk of COVID-19 related death, and those who survive are at risk of hospital readmission, post-discharge COVID-19 related mortality, and post-discharge COVID-19 related costs. The model includes risk of COVID-19 related death for hospitalized patients only; all-cause mortality and mortality related to COVID-19 in an outpatient care setting were not included in the model.

### COVID-19 INCIDENCE

The infection incidence estimates amongst the unvaccinated for January to December 2022 were based on projections from the Institute for Health Metrics and Evaluation (IHME)^23^ IHME provided information on the estimated number of infections amongst the unvaccinated as well as the number of people vaccinated in their simulation for 2022. The monthly rate of infections in the unvaccinated was calculated using this data. As the IHME model does not look at incidence by age, the same monthly rate is assumed for all age groups. Based on their modeling, IHME predicts that a large portion of individuals in countries such as the US have been infected with omicron meaning that immunity levels will be very high by March 2022. As shown in Table 2, the monthly rate of infection is projected to drop significantly after March.

**Table 2.** Monthly COVID-19 Incidence in unvaccinated individuals: Current and sensitivity analysis scenarios*.

|  | Current |  | Low |  | High |  |
| --- | --- | --- | --- | --- | --- | --- |
| Month | 18-64 yrs | ≥65 yrs | 18-64 yrs | ≥65 yrs | 18-64 yrs | ≥65 yrs |
| January | 32.38% | 32.38% | 25.90% | 25.90% | 38.85% | 38.85% |
| February | 6.21% | 6.21% | 4.97% | 4.97% | 7.45% | 7.45% |
| March | 2.03% | 2.03% | 1.62% | 1.62% | 2.43% | 2.43% |
| April | 0.71% | 0.71% | 0.57% | 0.57% | 0.86% | 0.86% |
| May | 0.33% | 0.33% | 0.26% | 0.26% | 0.40% | 0.40% |
| June | 0.16% | 0.16% | 0.13% | 0.13% | 0.19% | 0.19% |
| July | 0.10% | 0.10% | 0.08% | 0.08% | 0.11% | 0.11% |
| August | 0.08% | 0.08% | 0.06% | 0.06% | 0.09% | 0.09% |
| September | 0.09% | 0.09% | 0.07% | 0.07% | 0.11% | 0.11% |
| October | 0.20% | 0.20% | 0.16% | 0.16% | 0.24% | 0.24% |
| November | 0.63% | 0.63% | 0.50% | 0.50% | 0.75% | 0.75% |
| December | 2.33% | 2.33% | 1.86% | 1.86% | 2.79% | 2.79% |

\* January to December are based on projections from IHME

Low and high scenarios also were created by increasing and decreasing the base case cumulative incidence rates by 20% for January to December 2022. The final monthly incidence rates estimate for each scenario are displayed in Table 2. As the IHME projections count all infections including those that are asymptomatic, we reduced the estimate to 85% of the original to account for symptomatic cases only.^26^

### VACCINE EFFECTIVENESS

As the characteristics of the next variant that will become dominant in the US is unknown, the effectiveness of the existing vaccines to future variants is uncertain. Thus, three hypothetical scenarios, using existing variants as proxies, are explored:

1. Base case: A variant with initial VE similar to omicron: moderate initial VE from primary series and booster against infection and severe disease. Waning of VE is slow.
2. High effectiveness scenario: A variant with initial VE similar to delta: high initial VE from primary series and booster against infection and severe disease. Waning of VE is slow.
3. Low effectiveness scenario: A variant with initial VE similar to omicron with faster waning: moderate initial VE from primary series and booster against infection and severe disease. The rate of waning of VE is double that of the base case and the high effectiveness scenario.

The initial VE and rate of waning for each vaccine and each scenario is displayed in Table 3.

**Table 3.**
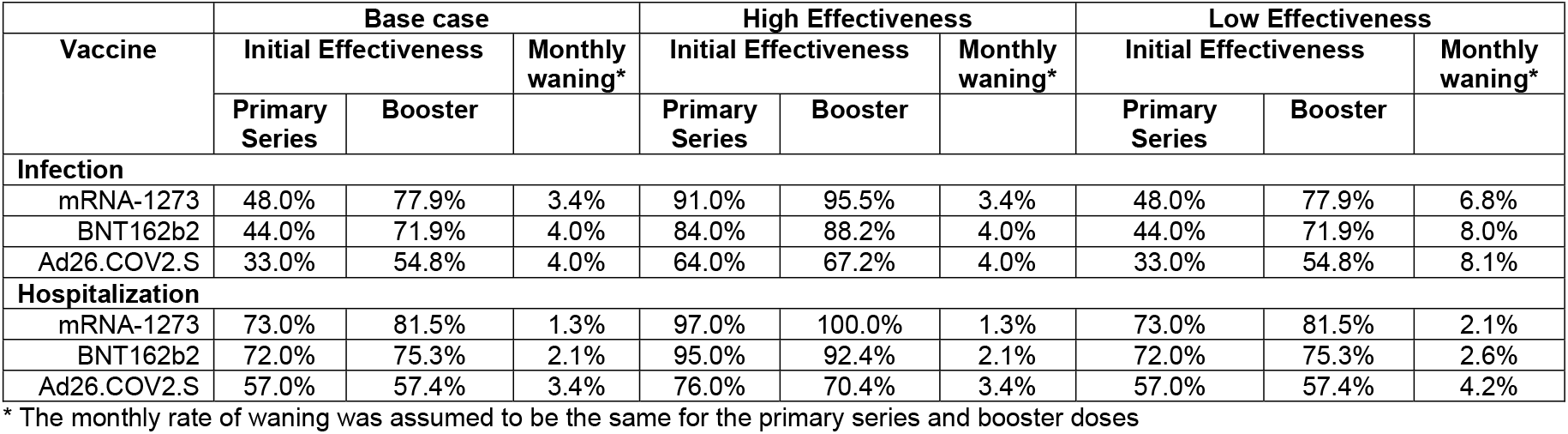
Vaccine Effectiveness Scenarios.

| Vaccine | Base case |  |  | High Effectiveness |  |  | Low Effectiveness |  |  |
| --- | --- | --- | --- | --- | --- | --- | --- | --- | --- |
|  | Initial Effectiveness |  | Monthly waning* | Initial Effectiveness |  | Monthly waning* | Initial Effectiveness |  | Monthly waning* |
|  | Primary Series | Booster |  | Primary Series | Booster |  | Primary Series | Booster |  |
| **Infection** |  |  |  |  |  |  |  |  |  |
| mRNA-1273 | 48.0% | 77.9% | 3.4% | 91.0% | 95.5% | 3.4% | 48.0% | 77.9% | 6.8% |
| BNT162b2 | 44.0% | 71.9% | 4.0% | 84.0% | 88.2% | 4.0% | 44.0% | 71.9% | 8.0% |
| Ad26.COV2.S | 33.0% | 54.8% | 4.0% | 64.0% | 67.2% | 4.0% | 33.0% | 54.8% | 8.1% |
| **Hospitalization** |  |  |  |  |  |  |  |  |  |
| mRNA-1273 | 73.0% | 81.5% | 1.3% | 97.0% | 100.0% | 1.3% | 73.0% | 81.5% | 2.1% |
| BNT162b2 | 72.0% | 75.3% | 2.1% | 95.0% | 92.4% | 2.1% | 72.0% | 75.3% | 2.6% |
| Ad26.COV2.S | 57.0% | 57.4% | 3.4% | 76.0% | 70.4% | 3.4% | 57.0% | 57.4% | 4.2% |

\* The monthly rate of waning was assumed to be the same for the primary series and booster doses

To inform the initial VE and the rate of waning over time following primary series for all VE scenarios, VE data for all 3 vaccines from the December 22, 2021 IHME COVID-19 model update estimates for delta and omicron were used, respectively.^11^ IHME pooled findings from multiple studies to calculate VE for the delta variant. They then calculated the relative reduction in VE against omicron compared to delta from two test-negative case control studies. This relative reduction was then applied to the delta VE to obtain the omicron VE. The monthly rates of waning for VE against infection and severe disease for each vaccine were approximated from the IHME graphs (timepoint 0-50 weeks). To create the graphs, IHME pooled data from 20 studies that estimated VE as a function of time, and used regression models to estimate the rate of decline. Waning from boosters were assumed to be the same as waning from primary series.

Booster VE against infection was not available from IHME. Therefore, the initial VE following booster vaccination for the high effectiveness scenario was estimated by applying the ratio of booster initial VE to primary series initial VE against delta infection estimated by Andrews et al., 2021 for BNT162b2 (1.05) to the IHME primary series initial VE for each vaccine. A similar ratio against omicron infection was not available from Andrews et al., 2021.^12^ However, the ratio of booster initial VE for omicron infection to booster initial VE for delta infection for BNT162b2 was estimated at 0.82. Therefore, to calculate the initial VE for infection following booster for the base case and low effectiveness scenario, this ratio was applied to the initial VE for infection following booster used in the high effectiveness scenario for each vaccine.

Booster VE against severe disease was not available from IHME or Andrews et al., 2021.^12^ Cromer et al., 2021^18^ estimated the ratio of booster VE against severe disease to booster VE against infection for delta at 1.05 for mRNA-1273. This ratio was applied to the booster VE for infection for all vaccines in the high effectiveness scenario to approximate the booster VE against severe disease.

Finally, to estimate the booster VE against severe disease for the base case and low effectiveness scenario, VE data from Andrews et al., 2021^12^ were used. The ratio of booster VE against infection for omicron to booster VE against infection for delta was estimated at 0.82 for BNT162b2. This was applied to the booster VE against severe disease from the high effectiveness scenario for each vaccine to calculate the base case and low effectiveness scenario values.

### INFECTION CONSEQUENCES

The age-specific proportions of symptomatic infections and patients requiring inpatient care were estimated from Reese et al., 2021 (see Appendix Table 6).^26^ The age-specific distributions of hospitalized patients across locations of care were estimated based on Di Fusco et al., 2021^27^ who performed a claims analysis using patients with a primary or secondary discharge diagnosis code for COVID-19 (ICD-10 code U07.1) from April 1 - October 31, 2020 in the Premier Healthcare COVID-19 Database. Age-specific estimates of in-hospital mortality were also calculated based on data provided by Di Fusco et al.^27^

Hospital re-admission rates were calculated based on data from Verna et al., 2021^28^ and are applied to all patients who survive their initial hospital stay in the model. Based on the highest level of oxygen received, patients were categorized into the various locations of care. Either ‘no oxygen’ or ‘low flow oxygen’ in the Verna study was assumed to correspond to ‘no ICU or ventilator’, ‘high flow’ was assumed to correspond to ‘ICU only’, and ‘ECMO’ or ‘mechanical ventilation’ was assumed to correspond to ICU with ‘ventilator’. The total number of patients re-admitted to the hospital in each location of care was divided by the total number of patients in each location of care. Verna et al., 2021^28^ did not provide data by age category; accordingly, the same percentages of patients requiring hospital re-admission within each location of care were applied to all age groups. See Appendix Table 6 for inputs used in the model.

For those patients surviving the initial hospitalization, the rates of post-discharge mortality were calculated from Chopra et al., 2021.^29^ Of those discharged, 1.96% were assumed to be readmitted to the ICU and require mechanical ventilation, 2.62% admitted to the ICU without ventilation, and 3.77% admitted to the hospital without ICU requirements.^28^ Additionally, the rates of post-hospital discharge mortality were assumed to be 10.40% for patients in ICU with/without ventilation, and 5.36% for those not requiring ICU admission.^29^ These rates were applied to all age groups.

### COSTS

Vaccine costs (including administration fees) were not included in the model, as they were assumed to be equivalent for all vaccine types and therefore do not contribute to the economic impact.

Symptomatic patients were assumed to receive one outpatient visit each. The proportion of patients requiring an emergency department visit without admission and the cost per outpatient visit were obtained from published sources.^26,30^ Inpatient costs were applied based on whether ICU or mechanical ventilation was required, and obtained from Di Fusco et al., 2021.^27^ Finally, Chopra et al., 2021^29^ estimated that 78.5% of patients that survived an inpatient stay required a primary care follow-up post-discharge. Unit costs for outpatient and inpatient care are provided in Appendix Table 8. Costs are expressed in 2022 USD; as costs are estimated over the period of one year only, discounting is not applied.

## 3. RESULTS

The number of outpatient visits, hospitalizations, and deaths and the medical costs associated with treating COVID-19 for each market share scenario is shown in Table 4 for the base case analysis. In the Current Scenario (mixed boosters strategy), the model predicts 65.2 million outpatient visits, 3.4 million hospitalizations, and 636,100 deaths from COVID-19 in 2022. Given the higher overall effectiveness for the mRNA-1273 booster compared to the BNT162b2 booster, if 100% of people received this booster, each of those outcomes would be expect to decrease by at least 1% compared to the Current Scenario. Specifically, 684,400 fewer outpatient visits, 48,700 fewer hospitalizations and 9,500 fewer deaths would be expected. As all vaccine costs are assumed to be the same, this is expected to decrease direct medical costs by $1.3 billion. Conversely, the BNT162b2 Scenario increased these outcomes compared to the Current Scenario. Specifically, 391,500 more outpatient visits, 34,500 more hospitalizations and 7,200 more deaths would be expected in 2022, increasing direct medical costs by $946 million. For both the mRNA-1273 and BNT162b2 booster scenarios, the percent change in direct treatment costs of COVID-19 is similar to the percent change in hospitalizations as the rate of hospitalizations is the driver of the overall costs.

**Table 4.** Base-Case Results.

| mRNA-1273 Scenario<br>(100% mRNA-1273) |  | Current Booster<br>Market Share | BNT162b2 Scenario<br>(100% BNT162b2) |  |
| --- | --- | --- | --- | --- |
| % Change from<br>Current | Change from<br>Current |  | Change from<br>Current | % Change from<br>Current |
| Healthcare Resource Utilization & Clinical Outcomes |  |  |  |  |
| Number of Outpatient Visits |  |  |  |  |
| -1.05% | -684,412 | 65,225,424 | 391,479 | 0.60% |
| Number of Hospitalizations |  |  |  |  |
| -1.44% | -48,662 | 3,378,170 | 34,454 | 1.02% |
| Number of Deaths |  |  |  |  |
| -1.49% | -9,487 | 636,099 | 7,217 | 1.13% |
| Total Costs, by Component and Overall (in millions) |  |  |  |  |
| Outpatient Visit Costs |  |  |  |  |
| -1.05% | -\$94 | \$8,949 | \$54 | 0.60% |
| Hospitalization Costs |  |  |  |  |
| -1.45% | -\$1,243 | \$85,755 | \$893 | 1.04% |
| Direct Medical Costs |  |  |  |  |
| -1.41% | -\$1,337 | \$94,704 | \$946 | 1.00% |

Given that January experienced a high incidence due to the omicron wave, we also looked at the results for February to December only, excluding January 2022 cases. There are fewer COVID-19 infections and therefore only 17.9 million outpatient visits, 965,600 hospitalizations, and 182,800 deaths. However, use of 100% mRNA-1273 still leads to a reduction in these outcomes by 2.3%-2.8%, while use of 100% BNT126b2 increases them by approximately 1.4%-2.1%.

Figure 2 shows the impact of changing incidence from the base case incidence to either the high or low incidence scenarios. The incidence impacts the total number of cases projected in 2022: higher incidence leads to higher outpatient visits, hospitalizations and deaths for all scenarios. With all incidence scenarios, additional use of mRNA-1273 leads to fewer infection outcomes while additional use of BNT126b2 leads to higher infection outcomes. The difference between the two vaccines, however, is more pronounced with higher infection incidence.

**Figure 2.**
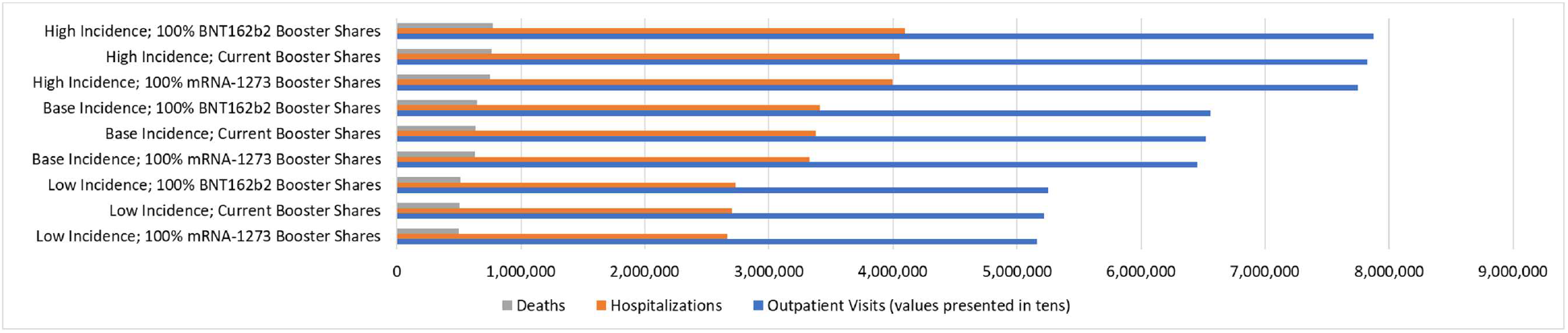
Clinical Outcomes, by Incidence and Market Share*. *All analyses performed assuming base-case effectiveness (100% Omicron)

The impact of varying the VE assumptions is displayed in Figure 3. All outcomes are increased when vaccine effectiveness is lower (omicron-like VE with double waning scenario) and decreased when VE is higher (delta-like VE scenario). In all scenarios, the mRNA-1273 booster prevents more cases than the BNT162b2 booster. Overall, the trends are as expected but the analysis quantifies the expected differences between the booster scenarios.

**Figure 3.**
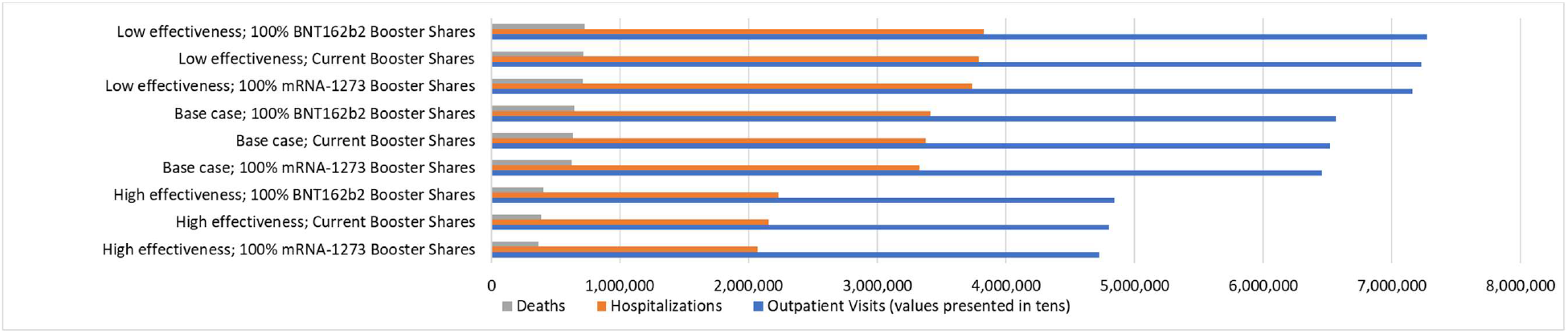
Clinical Outcomes, by Effectiveness and Market Share*. *All analyses performed using the base-case incidence

## 4. DISCUSSION

This study found that, when compared to the booster mix of both mRNA-1273 and BNT162b2 observed in the US as of December 2021, shifting to 100% mRNA-1273 boosters decreases the expected number of outpatient visits, hospitalization, and deaths, which in turn results in decreased direct medical costs. Conversely, shifting to boosting with 100% BNT162b2 increased resource use and medical costs. These trends remained consistent across a range of sensivitiy analyses varying the incidence of COVID-19 and the VE against different variant types.

The results are not surprising: as the predicted VE of the mRNA-1273 has been shown to be higher against infection and severe disease compared to BNT162b2 for existing variants,^11,13,18,21,31^ shifting to exclusive mRNA-1273 use as a booster would result in fewer COVID-19 cases and therefore less health care resource use.

As with all predictive modeling exercises, there are limitations to the study. Firstly, there is high uncertainty regarding the VE against emerging variants. Additionally, VE may increase as manufacturers tailor the vaccine to improve protection against new strains. Regardless of the variant, studies have shown higher VE for mRNA-1273 compared to BNT162b2, and therefore this relationship was maintained in our hypothetical VE profiles in the model for both primary and booster vaccinations. The base case analysis results assume VE similar to that estimated against an omicron-like variant. To address the uncertainty surrounding VE, different scenarios were tested. Assuming a high initial VE still resulted in the same trends as the base case; 100% mRNA-1273 boosters decreased, and 100% BNT162b2 boosters increased outpatient visits, hospitalization and deaths, compared to the booster mix. Assuming a low initial VE with double the monthly rate of waning compared to the base case also maintained the same trends as the base case. With lower overall effectiveness for all vaccines the number of cases and associated resource use is higher in all booster mix scenarios.

The incidence of COVID-19 had to be projected over the upcoming year and is highly uncertain. Still, scenarios where a high incidence rate and low incidence rate were tested did not change the overall model results, with 100% mRNA-1273 boosters resulting in less outpatient visits, hospitalizations and costs compared to the Current Scenario and 100% BNT162b2. With a higher incidence, VE is amplified as the overall denominator of cases is larger. Overall, resource use, deaths and costs are increased, and the difference in effects of booster types is also larger than the base case. An additional limitation of the incidence used is that the incidence data included in the model contains the rapid rise of the omicron wave in January 2022 and subsequent drop in February 2022. The January 2022 incidence (33%) contributed a large proportion of cases to the model. If COVID-19 becomes endemic, this high monthly incidence is not expected to occur again.^24^ However, the order of benefit (100% mRNA-1273 > booster mix > 100% BNT162b2) observed in the base case is still maintained for the February to December results only.

As the final proportion of people who choose to receive boosters is yet unknown, an assumption about future booster coverage had to be made in order to conduct this analysis. Demand for booster doses was very strong in January and February of this year as people sought additional protection against the new omicron variant. It is possible that 95% coverage will not be achieved if demand for the booster declines as incidence of disease declines. The final booster coverage achieved will impact the magnitude of cost-savings predicted: as more people are vaccinated, more individuals would receive benefit from a more-effective booster and this improves the cost savings associated with the mRNA-1273 scenario compared to the BNT162b2 scenario.

Some infectious disease models track susceptibility to infection in order to calculate an overall rate of infection in a population.^32^ We used a simplified model where the incidence rate amongst the unvaccinated is an input rather than a calculation in the model. In January to March, many people, vaccinated and unvaccinated are expected to be exposed to the omicron variant leading to increased levels of susceptibility. We indirectly account for that by using a lower incidence rate in the last 6 months of the year. However, one limitation is that we are not able to directly test the impact of different levels of immunity for individuals who receive a booster alone compared to those who receive a booster plus a COVID-19 infection.

The risk of hospitalization and the proportion of infections that were symptomatic in unvaccinated individuals were kept constant in all analyses. It is possible that emerging variants may differ in virulence than what is currently assumed in the model. The impact of increased booster coverage would be greater if the variant is more virulent. Additionally, this study reflects the clinical environment as of January 2022, a time at which there are currently limited treatment options available for COVID-19. Although oral antiviral pills have been approved for emergency use authorization (EUA) by the Food and Drug Administration (FDA),^33^ the impact on hospitalizations have not been included in the model.

Hospital re-admission data from Verna et al., 2021^28^ suggest a higher risk of re-hospitalization for less severe cases, perhaps as a result of increased in-hospital mortality for more severe cases or a discharge to hospice care for those severely affected by COVID-19. The current study assumed no risk of mortality for outpatient COVID cases, which may underestimate the true COVID-related mortality rate.

Adverse events associated with vaccines were not included in the model. There are limited data on whether injection-related and systemic adverse events are different between the vaccine types and serious adverse events are rare. For example, Oster et al., 2022^34^ report that of 192,405,448 people who received mRNA vaccines, 1,626 experienced myocarditis (<0.001%). While a single case of myocarditis may be expensive to diagnose and treat, the rarity of the outcome means that the occurrence of myocarditis has minimal impact on the costs. On the other hand, individuals who develop COVID-19 are at an increased risk of developing myocarditis^35^ and we did not consider the costs associated with that potential complication of COVID-19 either.

Finally, total costs savings are likely to be an underestimation of true values as many parameters, which can be significant drivers of cost, have not been included in the analyses. Productivity loss due to acute infection, or quarantine/isolation requirements due to infection/exposure have not been included in the model. Post-acute sequelae of COVID-19 (long Covid), which could impact both productivity loss as well as increase direct medical costs, has also not been incorporated into this model. It is expected that increasing booster vaccinations would decrease rates of infection and decrease these additional costs.

## 5. CONCLUSION

In conclusion, as the real-world effectiveness evidence to date indicates that mRNA-1273 may be more effective at preventing COVID-19 infection and hospitalization over time than BNT162b2, increasing the proportion of people receiving this as a booster may reduce the number of cases and costs associated with COVID-19 in 2022, regardless of COVID-19 incidence or variant.

## Data Availability

All data produced in the present work are contained in the manuscript.

## Author Contributions

AL, KF, MK, MM, PB and NV were involved in study design and interpretation of the analysis. MM programmed the model with quality assurance by MK, AL, and KF. All authors were involved in model estimation. MM and KF conducted the analysis. AL, KF and MK wrote the initial draft of the manuscript, and all remaining co-authors critically revised the manuscript and approved the final version.

## 6. APPENDIX

**Table 5.** Population Distribution, by Age Group.

| Age Group (yrs) | Population Size (N) | Source |
| --- | --- | --- |
| 18-29 | 53,728,222 | United States Census Bureau<sup>22</sup> |
| 30-39 | 44,168,826 |  |
| 40-49 | 40,319,374 |  |
| 50-64 | 62,925,688 |  |
| 65-74 | 31,483,433 |  |
| 75-84 | 15,969,872 |  |
| ≥85 | 6,604,958 |  |

**Table 6.** Percentage of Symptomatic vs. Asymptomatic Infections.

| Age Group (yrs) | Percentage Symptomatic | Percentage Asymptomatic | Source |
| --- | --- | --- | --- |
| 18-29 | 85.3% | 14.74% | Reese et al., 2021<sup>26</sup> |
| 30-39 | 85.3% | 14.74% |  |
| 40-49 | 85.3% | 14.74% |  |
| 50-64 | 85.3% | 14.74% |  |
| 65-74 | 80.8% | 19.16% |  |
| 75-84 | 80.8% | 19.16% |  |
| ≥85 | 80.8% | 19.16% |  |

**Table 7.** Hospitalization Inputs.

| Age (yrs) | % of Infections Requiring Hospitalizations<sup>26</sup> | Location of Care (Initial Admission)<sup>27</sup> |  |  | In-Hospital Mortality<sup>27</sup> |  |  |
| --- | --- | --- | --- | --- | --- | --- | --- |
|  |  | No ICU or Ventilator | ICU Only | Ventilator | No ICU or Ventilator | ICU Only | Ventilator |
| 18-29 | 2.60% | 86.86% | 7.23% | 5.91% | 0.02% | 0.56% | 18.94% |
| 30-39 | 2.60% | 82.39% | 8.59% | 9.03% | 0.10% | 0.68% | 26.18% |
| 40-49 | 2.60% | 77.17% | 9.31% | 13.52% | 0.20% | 0.90% | 34.72% |
| 50-64 | 7.20% | 71.27% | 9.61% | 19.12% | 1.13% | 3.57% | 46.36% |
| 65-74 | 22.44% | 67.41% | 9.80% | 22.80% | 4.25% | 10.38% | 58.68% |
| 75-84 | 22.44% | 70.40% | 10.04% | 19.56% | 10.15% | 22.58% | 66.94% |
| ≥85 | 22.44% | 79.37% | 8.93% | 11.70% | 19.69% | 32.37% | 72.47% |

**Table 8.**
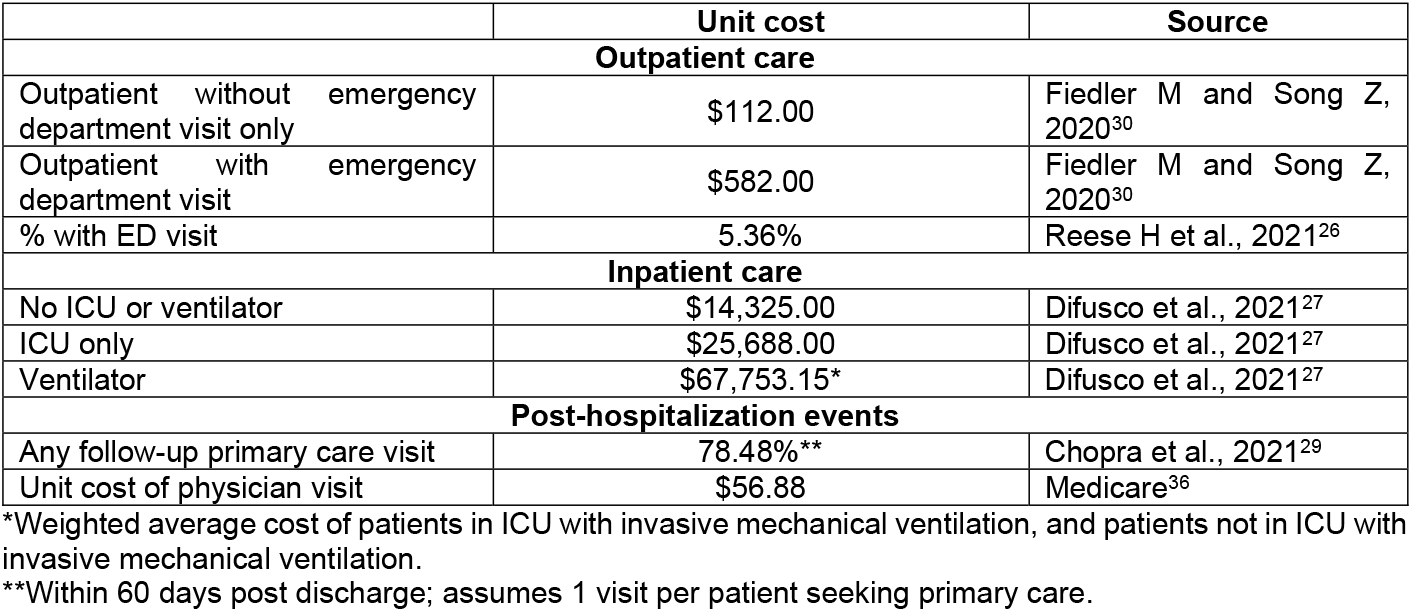
Cost Inputs.

|  | Unit cost | Source |
| --- | --- | --- |
| **Outpatient care** |  |  |
| Outpatient without emergency department visit only | \$112.00 | Fiedler M and Song Z, 2020<sup>30</sup> |
| Outpatient with emergency department visit | \$582.00 | Fiedler M and Song Z, 2020<sup>30</sup> |
| % with ED visit | 5.36% | Reese H et al., 2021<sup>26</sup> |
| **Inpatient care** |  |  |
| No ICU or ventilator | \$14,325.00 | Difusco et al., 2021<sup>27</sup> |
| ICU only | \$25,688.00 | Difusco et al., 2021<sup>27</sup> |
| Ventilator | \$67,753.15* | Difusco et al., 2021<sup>27</sup> |
| **Post-hospitalization events** |  |  |
| Any follow-up primary care visit | 78.48%** | Chopra et al., 2021<sup>29</sup> |
| Unit cost of physician visit | \$56.88 | Medicare<sup>36</sup> |

\*Weighted average cost of patients in ICU with invasive mechanical ventilation, and patients not in ICU with invasive mechanical ventilation.
\*\*Within 60 days post discharge; assumes 1 visit per patient seeking primary care.

## References

1. Mbaeyi S, Oliver SE, Collins JP, et al. The Advisory Committee on Immunization Practices’ Interim Recommendations for Additional Primary and Booster Doses of COVID-19 Vaccines - United States, 2021. MMWR Morbidity and mortality weekly report 2021; 70(44): 1545–52.

2. FDA U.S. Food & Drug Administration. Spikevax and Moderna COVID-19 Vaccine. Available at: https://www.fda.gov/emergency-preparedness-and-response/coronavirus-disease-2019-covid-19/spikevax-and-moderna-covid-19-vaccine. Accessed: February 16, 2022.; 2022(February 16).

3. FDA U.S. Food & Drug Administration. Comirnaty and Pfizer-BioNTech COVID-19 Vaccine. Available at: https://www.fda.gov/emergency-preparedness-and-response/coronavirus-disease-2019-covid-19/comirnaty-and-pfizer-biontech-covid-19-vaccine. Accessed: February 16, 2022.

4. FDA U.S. Food & Drug Administration. Janssen COVID-19 Vaccine. Available at: https://www.fda.gov/emergency-preparedness-and-response/coronavirus-disease-2019-covid-19/janssen-covid-19-vaccine. Accessed: February 16, 2022.

5. Centers for Disease Control and Prevention. Johnson and Johnson’s Janssen COVID-19 vaccine overview and safety. https://www.cdc.gov/coronavirus/2019-ncov/vaccines/different-vaccines/janssen.html Updated: December 28, 2021. Accessed: January 28, 2022.

6. Oliver SE, Wallace M, See I, et al. Use of the Janssen (Johnson & Johnson) COVID-19 Vaccine: Updated Interim Recommendations from the Advisory Committee on Immunization Practices - United States, December 2021. MMWR Morbidity and mortality weekly report 2022; 71(3): 90–5.

7. Centers for Disease Control and Prevention. COVID-19 Vaccine Booster Shots. Available at: https://www.cdc.gov/coronavirus/2019-ncov/vaccines/booster-shot.html. Updated: February 2, 2022. Accessed: February 4, 2022.

8. Centers for Disease Control and Prevention. COVID-19 Vaccine Emergency Use Instructions (EUI) Resources. Available at: https://www.cdc.gov/vaccines/covid-19/eui/index.html. Accessed: February 16, 2022.

9. Centers for Disease Control and Prevention. COVID Data Tracker. Available at: https://covid.cdc.gov/covid-data-tracker/#vaccination-demographic; Accessed: February 10, 2022.

10. Centers for Disease Control and Prevention. COVID Data Tracker. Available at: https://covid.cdc.gov/covid-data-tracker/#vaccination-demographic; Accessed: December 9, 2021.

11. Institute for Health Metrics and Evaluation (IHME). COVID-19 model update: Omicron and waning immunity. Available at: www.w.healthdata.org. Updated: December 22, 2021. Accessed: December 23, 2021.

12. Andrews N, Stowe J, Kirsebom F, et al. Effectiveness of COVID-19 vaccines against the Omicron (B.1.1.529) variant of concern. medRxiv 2021: 2021.12.14.21267615.

13. Tan S, Pung R, Wang L, et al. Protection of Homologous and Heterologous Vaccine Boosters Against COVID-19 in Singapore (November 16, 2021). Available at: https://ssrn.com/abstract=3995101 or http://dx.doi.org/10.2139/ssrn.3995101. Accessed: January 11, 2022.

14. Tseng HF, Ackerson BK, Luo Y, et al. Effectiveness of mRNA-1273 against SARS-CoV-2 omicron and delta variants. medRxiv 2022: 2022.01.07.22268919.

15. Drawz PE, DeSilva M, Bodurtha P, et al. Effectiveness of BNT162b2 and mRNA-1273 Second Doses and Boosters for SARS-CoV-2 infection and SARS-CoV-2 Related Hospitalizations: A Statewide Report from the Minnesota Electronic Health Record Consortium. medRxiv 2022: 2021.12.23.21267853.

16. Jackson ML, Nelson JC. The test-negative design for estimating influenza vaccine effectiveness. Vaccine 2013; 31(17): 2165–8.

17. Khoury DS, Cromer D, Reynaldi A, et al. Neutralizing antibody levels are highly predictive of immune protection from symptomatic SARS-CoV-2 infection. Nature medicine 2021; 27(7): 1205–11.

18. Cromer D, Steain M, Reynaldi A, et al. Neutralising antibody titres as predictors of protection against SARS-CoV-2 variants and the impact of boosting: a meta-analysis. Lancet Microbe 2022; 3(1): e52–e61.

19. Khoury DS, Steain M, Triccas JA, Sigal A, Davenport MP, Cromer D. A meta-analysis of Early Results to predict Vaccine efficacy against Omicron. medRxiv 2021: 2021.12.13.21267748.

20. Andrews N, Tessier E, Stowe J, et al. Vaccine effectiveness and duration of protection of Comirnaty, Vaxzevria and Spikevax against mild and severe COVID-19 in the UK. medRxiv 2021: 2021.09.15.21263583.

21. UK Health Security Agency. COVID-19 vaccine surveillance report. Week 4. January 27, 2022. Available at: https://assets.publishing.service.gov.uk/government/uploads/system/uploads/attachment_data/file/1050721/Vaccine-surveillance-report-week-4.pdf. Accessed: January 27, 2022.

22. U.S. Census Bureau. Population Division: Washington DC. Annual Estimates of the Resident Population by Single Year of Age and Sex for the United States: April 1, 2010 to July 1, 2019 (NC-EST2019-AGESEX-RES). Available at: https://www2.census.gov/programs-surveys/popest/technical-documentation/file-layouts/2010-2019/nc-est2019-agesex-res.csv. Accessed: November 3, 2021.

23. Institue for Health Metrics and Evaluation (IHME). COVID-19 Projections (January 27, 2022). United States of America. Used with permission. All rights reserved.

24. Murray CJL. COVID-19 will continue but the end of the pandemic is near. Lancet 2022; 399(10323): 417–9.

25. Molinari NA, Ortega-Sanchez IR, Messonnier ML, et al. The annual impact of seasonal influenza in the US: measuring disease burden and costs. Vaccine 2007; 25(27): 5086–96.

26. Reese H, Iuliano AD, Patel NN, et al. Estimated Incidence of Coronavirus Disease 2019 (COVID-19) Illness and Hospitalization-United States, February-September 2020. Clin Infect Dis 2021; 72(12): e1010–e7.

27. Di Fusco M, Shea KM, Lin J, et al. Health outcomes and economic burden of hospitalized COVID-19 patients in the United States. J Med Econ 2021; 24(1): 308–17.

28. Verna EC, Landis C, Brown RS, Jr., et al. Factors Associated with Readmission in the US Following Hospitalization with COVID-19. Clin Infect Dis 2021.

29. Chopra V, Flanders SA, O’Malley M, Malani AN, Prescott HC. Sixty-Day Outcomes Among Patients Hospitalized With COVID-19. Ann Intern Med 2021; 174(4): 576–8.

30. Fiedler M, Song Z. Brookings report: estimating potential spending on COVID-19 care. https://www.brookings.edu/research/estimating-potential-spending-on-covid-19-care/. Accessed: July 22, 2020.

31. Premika M, Chiew CJ, Wei WE, et al. Comparative Effectiveness of mRNA and Inactivated Whole Virus Vaccines against COVID-19 Infection and Severe Disease in Singapore (December 28, 2021). Available at: https://ssrn.com/abstract=3995282 or http://dx.doi.org/10.2139/ssrn.3995282. Accessed: January 11, 2022.

32. Pitman R, Fisman D, Zaric GS, et al. Dynamic transmission modeling: a report of the ISPOR-SMDM Modeling Good Research Practices Task Force--5. Value Health 2012; 15(6): 828–34.

33. Nadeem D, O’Donnell C. U.S. authorizes Pfizer oral COVID-19 treatment, first for at-home use. Available at: https://www.reuters.com/business/healthcare-pharmaceuticals/pfizer-oral-covid-19-pill-gets-us-authorization-at-home-use-2021-12-22/. Accessed: February 1, 2021.

34. Oster ME, Shay DK, Su JR, et al. Myocarditis Cases Reported After mRNA-Based COVID-19 Vaccination in the US From December 2020 to August 2021. JAMA 2022; 327(4): 331–40.

35. Barda N, Dagan N, Ben-Shlomo Y, et al. Safety of the BNT162b2 mRNA Covid-19 Vaccine in a Nationwide Setting. N Engl J Med 2021; 385(12): 1078–90.

36. Centers for Medicare and Medicaid Services. 2021 National Physician Fee Schedule Relative Value File October Release. Available at: https://www.cms.gov/Medicare/Medicare-Fee-for-Service-Payment/PhysicianFeeSched/PFS-Relative-Value-Files. Accessed: December 7, 2021.

